# Non-communicable disease and the risk of infection death: a UK Biobank prospective cohort study

**DOI:** 10.1101/2020.07.27.20162354

**Authors:** Michael Drozd, Patrick J Lillie, Mar Pujades-Rodriguez, Sam Straw, Ann W Morgan, Mark T Kearney, Klaus K Witte, Richard M Cubbon

**Affiliations:** Leeds Institute of Cardiovascular and Metabolic Medicine, The University of Leeds, Leeds, United Kingdom; Department of Infection, Castle Hill Hospital, Hull University Hospitals NHS Trust, Kingston Upon Hull, United Kingdom; Leeds Institute of Health Sciences, School of Medicine, University of Leeds, Leeds United Kingdom; NIHR Leeds Biomedical Research Centre, Leeds Teaching Hospitals NHS Trust, United Kingdom

## Abstract

**Objectives:** Non-communicable diseases (NCDs) have recently been highlighted as important risk factors for COVID-19 fatality. We set out to investigate the association between NCDs and the risk of death from any infection in the pre-COVID-19 era.

**Design:** Prospective population-based study

**Setting:** UK Biobank

**Participants:** 493,295 participants

**Main outcome measures:** Infection death prior to December 31^st^ 2019.

**Results:** During 5,277,344 participant-years of follow-up, 1,385 infection deaths occurred, accounting for 5% of all deaths. Competing risks regression revealed that advancing age, male sex, smoking, socio-economic deprivation and all studied NCDs were independently associated with both the risk of infection death and non-infection death; ethnicity was associated with neither. Only smoking, socio-economic deprivation, hypertension, respiratory disease, chronic kidney disease, psychiatric disease and rheumatological disease were associated with greater hazard ratios for infection than non-infection death. Accrual of multimorbidity was also associated with a greater increases in the risk of infection death (HR 9.03 [95% confidence interval 6.61 to 12.34] for 5+ comorbidities versus none; p<0.001), than non-infection death (HR 5.68 [95% confidence interval 5.22 to 6.17] for 5+ comorbidities versus none; p<0.001).

**Conclusions:** Diverse NCDs are associated with increased risk of infection death, suggesting that recently reported associations with COVID-19 death may be non-specific. Moreover, only a subset of NCDs, together with the accrual of multimorbidity, smoking and socio-economic deprivation, are associated with greater relative risks of infection death than other causes of death. Further research is needed to define why these risk factors are biased toward infection death so that more effective preventative strategies can be targeted to high-risk groups.

**What is already known on this topic:**

- Infection contributes to approximately one in five deaths globally and this often occurs in people with non-communicable diseases (NCDs).
- Many NCDs have recently been highlighted as risk factors for fatal COVID-19 infection.

**What this study adds:**

- Whilst diverse NCDs are associated with greater risk of infection death, only some pose greater hazards of infection than non-infection death.
- The association of many NCDs with fatal COVID-19 infection is unlikely to be specific to this pathogen.

## Introduction

Non-communicable diseases (NCDs) have recently been highlighted as important risk factors for fatal COVID-19 infection,(1,2) prompting hypotheses that the virus specifically interacts with some comorbidities.(3–5) However, the wider context of infectious diseases in people with NCDs is often neglected in such proposals, in spite of this being much more common and potentially offering insight into the conserved and the disease-specific risks of COVID-19. The Global Burden of Disease Study revealed 11 million sepsis-related deaths during 2017, amounting to almost 20% of all deaths.(6) Sepsis-related death was more common in men, the elderly (and infants), and socio-economically deprived populations. Moreover, around 40% of cases had an NCD listed as the underlying cause of death, with cardiovascular diseases, diabetes, liver disease, COPD and kidney disease all being commonly implicated. Hence, there are clear parallels between the risk factors for sepsis death and COVID-19 death. In spite of a 30% reduction in global sepsis-related deaths between 1990 and 2017,(6) the issue of life-threatening infection in people with NCDs is likely to become more important in the context of globally increasing rates of multimorbidity and anti-microbial resistance.(7,8) Moreover, as populations become more affluent, NCDs are proportionally more common as an underlying cause of sepsis death,(6) underlining the growing relevance to many ‘economically developing’ populations. In spite of the scale and impact of this issue, little prospective research is published about the independent risks posed by specific NCDs, and multimorbidity, for infection-related death. To address this, we analysed the well-characterised UK Biobank cohort study using follow-up data prior to the COVID-19 pandemic.

## Methods

### Study design and data collection

The UK Biobank (UKB) cohort is a population-based prospective study that consists of 502,505 people aged between 37-73 years. Participants were recruited between 2006 and 2010, and attended one of 22 assessment centres across England, Scotland and Wales. Baseline biological measurements were recorded, and participants completed a touchscreen and nurse-led questionnaire, as described elsewhere.(9) UKB received ethical approval from the NHS Research Ethics Service (11/NW/0382); we conducted this analysis under application number 59585. All participants provided written informed consent. There was no patient and public involvement in the planning of this analysis.

### Assessment of demographic factors and morbidity

Age, sex, ethnicity and socioeconomic status were considered as potential risk factors. Age at recruitment was categorised into the following groups: <45, 45 to <50, 50 to <55, 55 to <59, 60 to <64 and 65+. Ethnicity was participant-classified within UKB-defined categories of white, mixed, Asian or British Asian, black or British black, Chinese or other ethnic group. Self-reported smoking status was reported as never, previous or current. Socioeconomic deprivation (SED) was classified into quintiles using the Townsend score. Obesity was classified using the World Health Organisation’s categorisation according to body mass index: class 1 (30.0 - 34.9 kg/m^2^), class 2 (35.0 -39.9), class 3 (≥40). Self-reported medical conditions recorded at study recruitment during face-to-face interview with a nurse were used to classify morbidities into groups (described in **Supplemental Table 1**). We selected a range of morbidities that represent a broad spectrum of common disease groups, including: hypertension, chronic heart disease (ischaemic heart disease and heart failure), chronic respiratory disease, diabetes, prior cancer, chronic liver disease, chronic kidney disease, prior stroke/TIA, other neurological disease, psychiatric disease and rheumatological disease (chronic inflammatory and autoimmune rheumatic disease, not non-inflammatory disorders).(10) The number of these comorbidities was calculated for each participant. Missing data for comorbidities (n=863), body mass index (n=3,106), smoking status (n=2,949), ethnicity (n=2,777) and SED (624) resulted in exclusion of those participants from the analysis.

### Mortality ascertainment

Mortality information was recorded through linkage to national death registries. In the present analysis, we censored deaths up until 31^st^ December 2019 to ensure this was before the first reported case of COVID-19 in the United Kingdom.(11) Deaths were classified using ICD-10 codes for the main cause of death as infection-related (described in **Supplemental Table 2**) or non-infection. Loss to follow-up or withdrawal of consent resulted in 863 participants being excluded from the analysis.

### Statistical analysis

Continuous variables are presented as mean (standard deviation) and categorical variables as number (percentage). Adjusted cause-specific mortality hazard ratios were estimated using Fine and Gray competing risk regression modelling,(12) with infection deaths as the events of interest and non-infection deaths as competing events, or *vice versa*, with the same adjustment variables. Models were adjusted for all covariates, including: age, sex, SED, smoking, obesity, hypertension, chronic heart disease, chronic respiratory disease, diabetes, cancer, chronic liver disease, chronic kidney disease, prior stroke/TIA, other neurological disease, psychiatric disorder and rheumatological disease (or the number of comorbidities, up to a maximum of 5, in place of obesity, hypertension, chronic heart disease, chronic respiratory disease, diabetes, cancer, chronic liver disease, chronic kidney disease, prior stroke/TIA, other neurological disease, psychiatric disorder and rheumatological disease). All tests were 2-sided and statistical significance was defined as p<0.05. All statistical analyses were performed using Stata (StataCorp LP, College Station, USA; version 16.1) and R (http://cran.r-project.org/; version 4.0.2).

## Results

Amongst the UK Biobank cohort of 502,505 people, we excluded 9,210 (1.8%) due to missing baseline data or long-term follow-up data, as described in the methods section. Within the study sample of 493,295 people, 1,385 infection deaths occurred during 5,277,344 person-years of follow-up (10.9, IQR 10.1 - 11.6), accounting for approximately 5% of the total 27,729 deaths (**Figure 1a**). The commonest fatal infections involved the lower respiratory tract (60.7%), gastrointestinal tract (14.2%) and genitourinary tract (5.9%); data for all causes are presented in **Figure 1b** (and **Supplemental Table 3**). Compared to survivors, people experiencing infection death were older, more often male, more socio-economically deprived, more commonly smoked, and were more likely to have a wide range of NCDs (**Table 1**). Broadly similar differences were observed when comparing survivors with people experiencing non-infection death, although cancer was markedly more common in people experiencing non-infection death.

**Figure 1:**
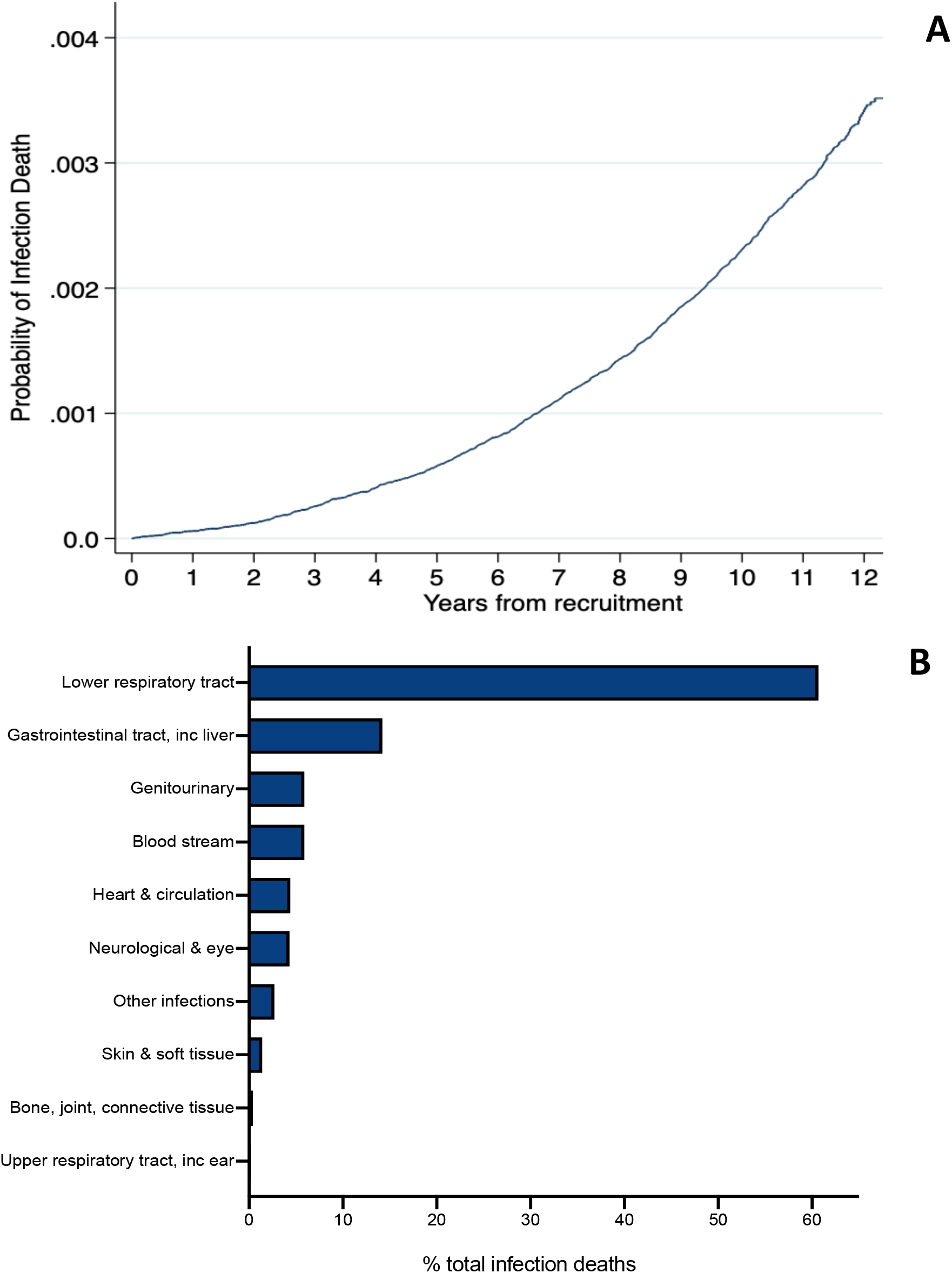
Timing and classification of infection deaths. **A**) Cumulative incidence function illustrating cumulative infection deaths during follow-up. **B**) Bar chart illustrating classification of infection deaths.

**Table 1:**
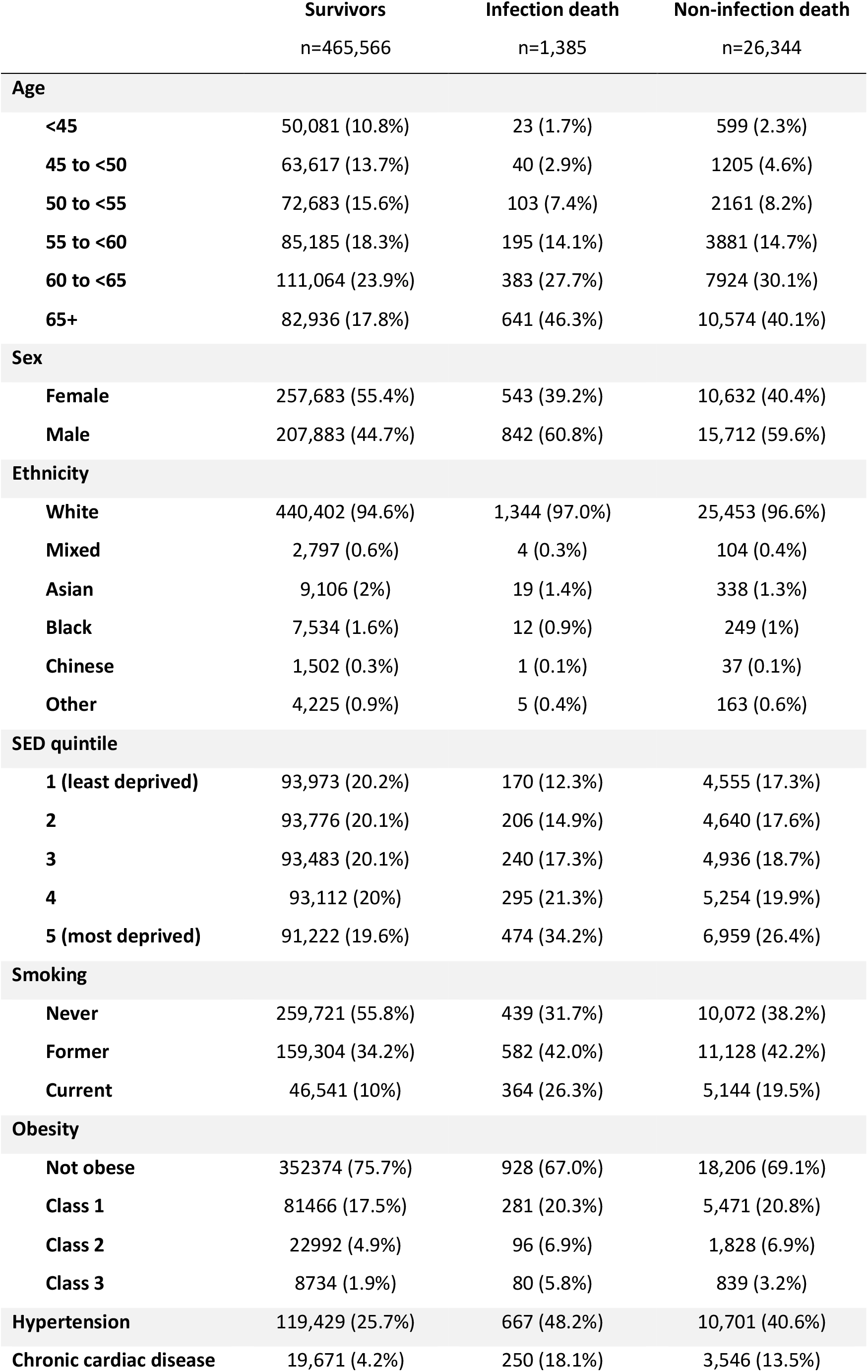
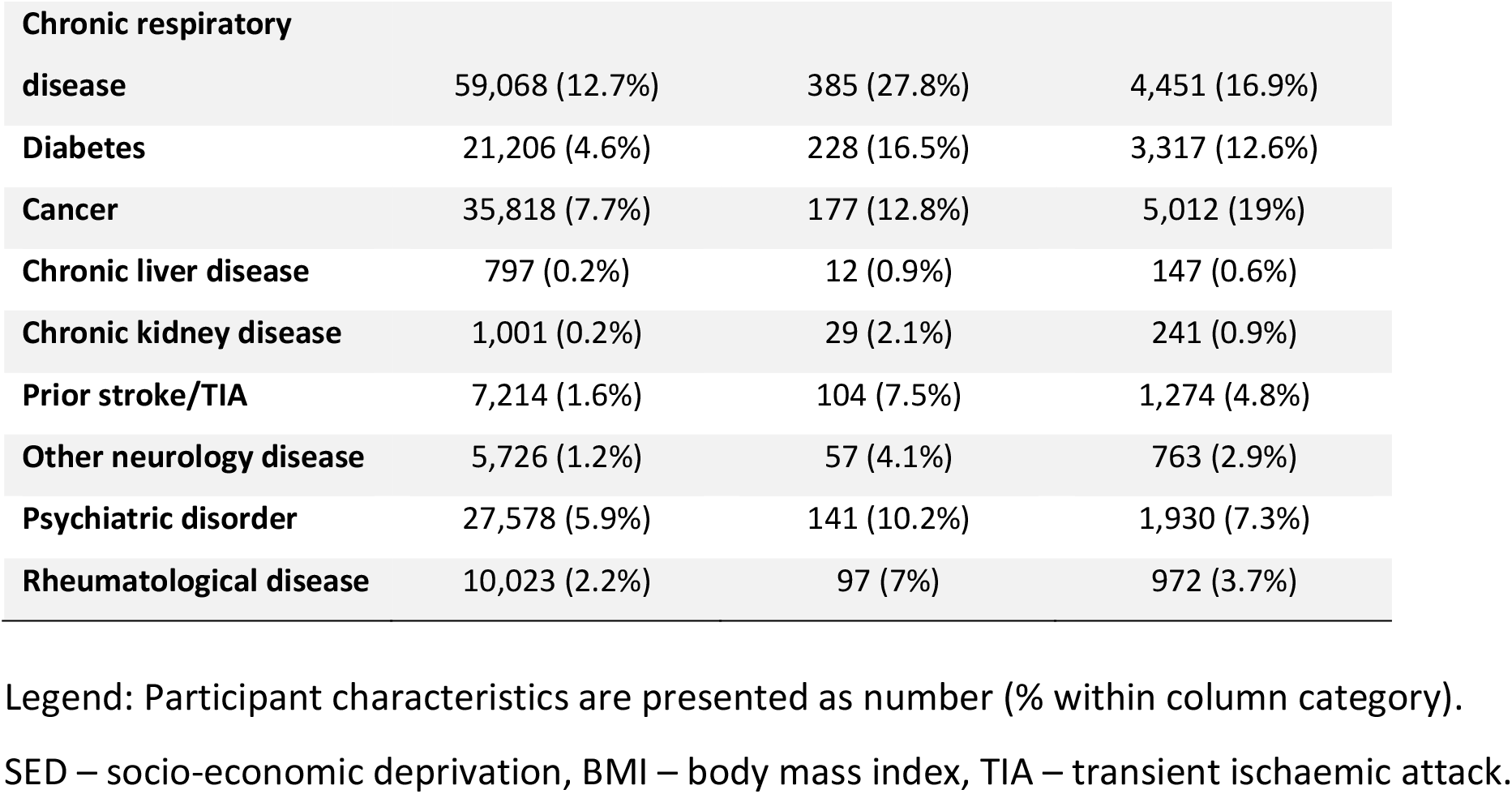
Participant characteristics at recruitment.

To define the association between NCDs and infection-death, whilst also accounting for the loss of participants due to non-infection death, we performed competing risks analyses to derive cause-specific hazard ratios. As shown in **Figure 2** (and **Supplemental Table 4**), advancing age, male sex, advanced obesity, smoking, SED and all individual NCDs were independently associated with the risk of infection death. However, there was no clear association between any ethnic group and altered risk of infection death. Similarly, for non-infection death, advancing age, male sex, obesity, smoking, socio-economic deprivation and all individual NCDs were also independently associated with the risk of infection death. Again, there was no clear association between any ethnic group and altered risk of non-infection death. However, the hazards associated with smoking, socio-economic deprivation, class 3 obesity, hypertension, chronic respiratory disease, chronic kidney disease, psychiatric disease and rheumatological disease were greater for infection than non-infection death, whilst the converse was apparent for cancer.

**Figure 2:**
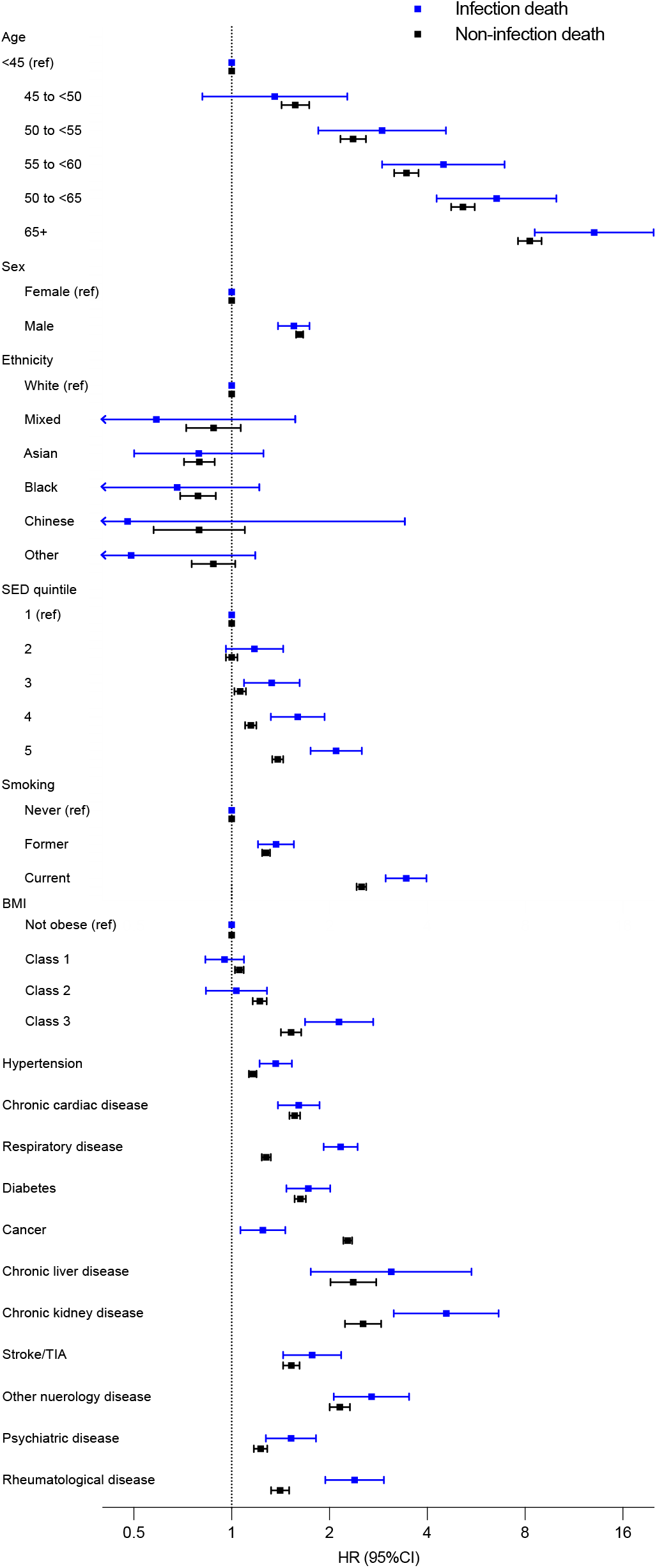
Association between participant characteristics and the risk of infection or non-infection death. Forest plot illustrating hazard ratios and 95% confidence intervals for infection (blue) or non-infection (black) death in multivariate competing risks regression analysis.

To explore the impact of multimorbidity, we conducted alternate competing risks analyses including the number of NCDs, rather than individual diseases, whilst also accounting for age, sex, ethnicity and socio-economic deprivation. As shown in **Figure 3** (and **Supplemental Table 5**), this revealed a clear gradient of increasing risk of infection death as the number of NCDs increased. Notably, whilst gradient of increasing risk of non-infection death was also observed, this was less steep than for infection death.

**Figure 3:**
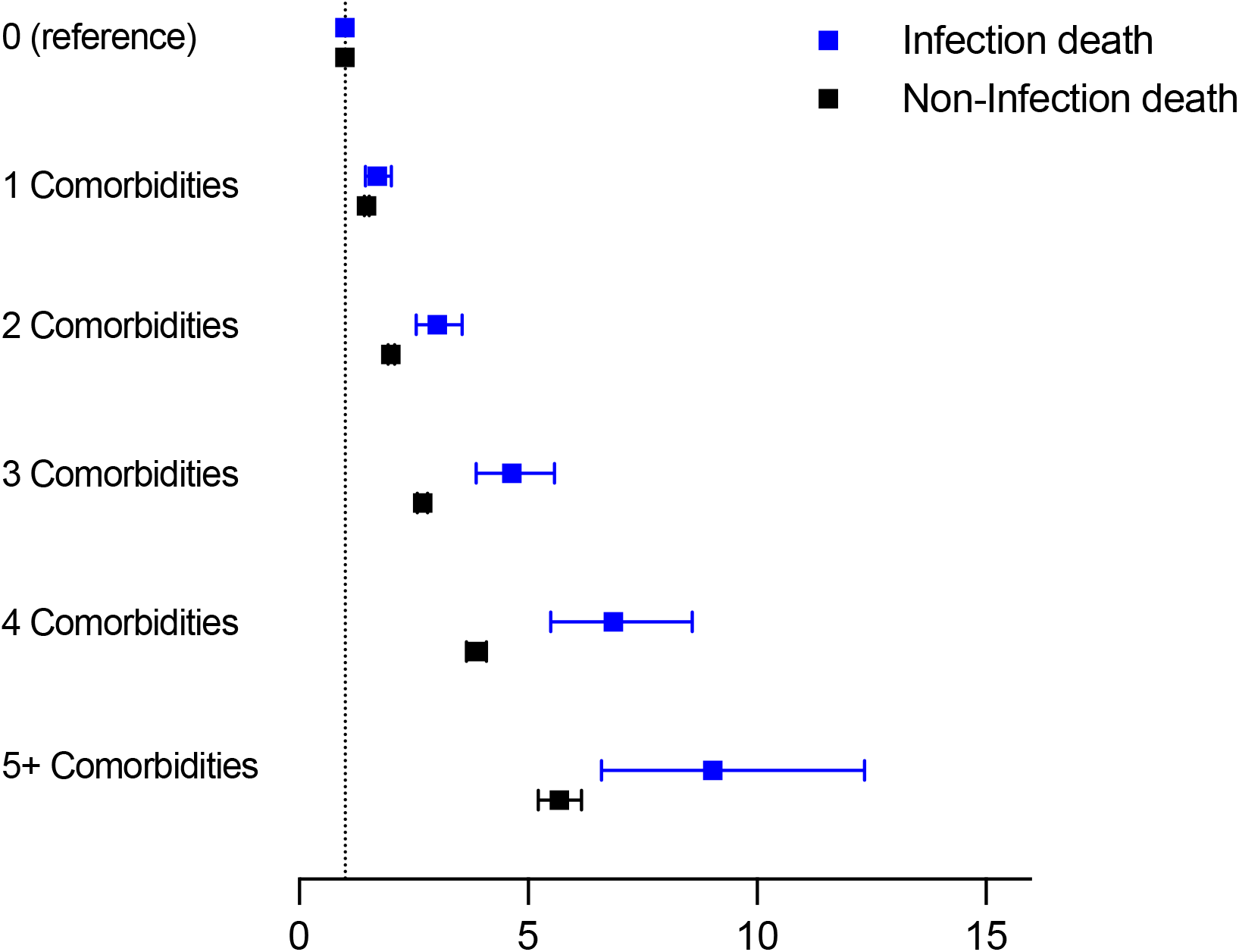
Association between multimorbidity and the risk of infection or non-infection death. Forest plot illustrating hazard ratios and 95% confidence intervals for infection (blue) or non-infection (black) death in multivariate competing risks regression analysis, adjusting for age, sex, socio-economic deprivation and ethnicity.

## Discussion

We have conducted the largest ever prospective analysis of risk factors for infection death in people recruited in a community setting. Infection death was more common in men, with advancing age and increasing SED. All studied NCDs were also independently associated with increased risk of infection death, as was the accrual of multimorbidity. Risk factors for infection death were similar to those reported for COVID-19 death, although most of these were associated with similar hazards of infection and non-infection death. However, some conferred greater risks of infection death, including: advancing age, smoking, socio-economic deprivation, class 3 obesity, hypertension, chronic respiratory disease, chronic kidney disease, psychiatric disease, rheumatological disease, and the accrual of multimorbidity. These high-risk populations may benefit from strategies to prevent or more effectively treat infection.

The existing literature supports the importance of infection as a cause of death in populations with many of the individual NCDs we have studied, including people with chronic heart failure;(13,14) diabetes,(15) chronic kidney disease,(16) liver disease,(17) rheumatological disease(18), and neurodegenerative disorders (19) Our analysis supports these disease-specific data, but has the advantage of considering both the additive and independent risk of infection death attributable to multiple NCDs in the same population. Notably, whilst the accumulation of multimorbidity has been linked with greater risks of all-cause, cardiovascular and cancer death,(10) our finding regarding a greater risk of infection than non-infection death are novel. Regarding the larger hazard of infection death associated with SED, whilst residual confounding could explain this association, socio-economic interventions have played major roles in addressing past and present infectious diseases.(20,21)

The COVID-19 pandemic has brought fresh attention to the adverse outcomes experienced by people with NCDs who develop infectious diseases. Whilst there are some disagreements regarding specific risk factors, probably reflecting differing methods, the COVID-19 literature broadly suggests that ageing, male sex, socio-economic deprivation, and accrual of NCDs increase the risk of adverse outcomes.(1,22) The broad agreement with our analysis of risk factors for death related to infection in the pre-COVID-19 era suggests common underlying factors and solutions to these problems. One notable difference is that ethnicity was not associated with infection death in our adjusted analyses, but has frequently been identified as a risk factor for adverse COVID-19 outcomes,(23) including in the UK Biobank cohort,(24) although not all studies agree.(1) Again, this disagreement may relate to methodological issues, but could also support COVID-19 differentially impacting specific ethnic groups.

It is important to acknowledge the potential limitations of our work. Firstly, UK Biobank is not a nationally representative cohort,(25) so caution should be applied in extrapolating data on infection death prevalence. However, our finding that infections account for 5% of all deaths is broadly in keeping with the Global Burden of Disease data on sepsis-death in the United Kingdom.(6) It is also important to highlight the challenges of death certification data, which do not always encompass the complex factors contributing to death. Similarly, the classification of NCD status in UK Biobank often relies on participant-reported data, and so may lack the fidelity of other approaches, although available adjudication data suggest these are valid.(26) Moreover, the classification of most NCDs as being present or absent neglects the heterogeneity within these, which may have an important influence on risk of infection-death. Finally, as with any observational study, it is important not to draw causal inferences from our data, although our findings raise many important and testable hypotheses.

Ongoing work is needed to understand the relative importance of NCDs causing recurrent infections, versus increasing the risk of death during any single episode, to the overall risk of infection death. Future research should also define mechanisms linking NCDs (and their treatment) to infection death e.g. via altered immune responses to vaccination and infection, inflammatory responses to pathogens, and physiological responses during infection/sepsis. Addressing these uncertainties will help to inform the selection of therapeutic strategies for clinical trials; these could include: specific primary and/or secondary infection prevention measures; improved access to care, monitoring for early detection of infection and initiation of appropriate antimicrobial treatment; improved use of high dependency care. However, even without further research, our data are important to health care professionals and people with NCDs, who should be alert to the importance of infection to adverse outcomes.

## Data Availability

The UK Biobank dataset is available to all bona fide researchers for all types of health-related research which is in the public interest.

## Acknowledgements

This research has been conducted using the UK Biobank Resource. MD is supported by a British Heart Foundation Clinical Research Training Fellowship. AM has received salary support from the National Institute for Health Research and the UK Medical Research Council. MTK is a British Heart Foundation professor. RMC was supported by a British Heart Foundation Intermediate Clinical Research Fellowship.

## Transparency Statement

RC affirms that the manuscript is an honest, accurate, and transparent account of the study being reported; no important aspects of the study have been omitted; there are no discrepancies from the study as originally planned.

## Contributorship Statement

Guarantors – MD and RC; Study conception – MD and RC; Data analysis – MD; Manuscript drafting – MD and RC; Critical revision of manuscript: PJL, MPR, AWM, SS, MTK, KKW. The corresponding author attests that all listed authors meet authorship criteria and that no others meeting the criteria have been omitted.

